# Sex differences in frailty trajectories among older adults in Mexico: a 17-year longitudinal cohort study

**DOI:** 10.64898/2026.06.25.26356559

**Authors:** Aleixo P Brunetti, Jennifer M Nicholas, Kwabena Asare, Kathryn E. Mansfield, Charlotte Warren-Gash

## Abstract

**Introduction:** Frailty is an ageing-related state associated with disability and mortality. Women often experience higher frailty but lower mortality than men, a pattern described as the male-female health-survival paradox. Evidence from low- and middle-income settings is limited. We examined sex differences in frailty trajectories and terminal decline in Mexico.

**Methods:** We analysed five waves (2001-2018) of the nationally representative Mexican Health and Aging Study (MHAS) including 12,440 adults (≥50 years at baseline). Frailty was measured using a 31-deficit frailty index (FI score; 0-1). We used survey-weighted linear mixed-effects models with time interactions, adjusted for sociodemographic, behavioural and health covariates to model sex differences in frailty trajectories. Terminal decline in FI was modelled among those who died using mixed-effects models on the time-to-death scale.

**Results:** A total of 12,440 adults aged 50 to 105 years were included, with a mean age of 62.1 years (SD 9.6); 5,698 men (45.8%) and 6,742 women (54.2%). Mean baseline FI was 0.17 (SD 0.12), higher in women than men (0.19 vs 0.16; P<0.001). After adjusting, women had a 0.014 higher mean FI than men at baseline (adjusted mean difference; 95%CI 0.008, 0.020), with difference widening over follow-up, increasing from 0.016 at 2 years to 0.029 at 17 years. Analysis of terminal decline found that accumulation of frailty accelerated in the years preceding death; with results suggesting that women reached death with higher frailty than men (difference 0.029; 95%CI 0.009, 0.048).

**Conclusion:** Women experienced higher and more rapidly increasing frailty compared to men and carried a greater frailty burden in the years preceding death. These findings underscore the importance of considering sex differences in frailty trajectories when developing healthy ageing strategies that address the life-course vulnerabilities disproportionately driving frailty accumulation in women in low- and middle-income countries.

## INTRODUCTION

Frailty is increasingly recognised as a major global health challenge as populations age.^1^ Frailty is characterised by diminished physiological reserve and heightened vulnerability to stressors, and is associated with reduced physical function, hospitalisation, and premature mortality.^2 3^ Although closely linked to ageing, frailty is not an inevitable consequence of older age; it is a dynamic and potentially reversible state that can worsen, stabilise, or improve over time.^3^ ^4^ Globally, 13.6% of community-dwelling older adults developed frailty on an average follow-up of just over 3 years, equivalent to 43.4 per 1,000 person-years, with substantially higher rates among prefrail individuals and in low- and middle-income countries (LMICs).^2 5 6^

Central to understanding frailty is the role of sex.^7^ Women consistently accumulate more health deficits than men yet survive longer (a pattern known as the male-female health-survival paradox) and are more likely to transition to frailty while men more often remain robust.^7^ ^8 9^ Despite their higher frailty burden, women experience lower mortality than men, and this survival advantage persists even after accounting for greater deficit accumulation.^7^ ^10^ In the years immediately before death, frailty accelerates sharply in both sexes, a phenomenon termed terminal decline, but evidence from high-income countries suggests this process begins earlier in women while being steeper in men.^6^

Most evidence on these sex differences originates from high-income countries, leaving fundamental questions unanswered in LMIC contexts.^7^ ^8^ ^11^ ^12^ Latin America and the Caribbean are among the world’s most rapidly ageing regions, where population ageing is unfolding alongside pronounced social inequalities and limited health and social protection systems, yet whether the health-survival paradox and sex differences in terminal decline operate similarly in these settings remains unknown.^13^ ^14^ Addressing this gap is essential for informing equitable, evidence-based strategies to promote healthy ageing in LMIC settings.

Therefore, our study aimed to investigate sex differences in frailty trajectories over 17 years of follow-up and sex differences in frailty trajectories in the years preceding death among older adults in Mexico.

## METHODS

### Study design, data source and population

We conducted a longitudinal cohort study using five waves (2001-2018) of the Mexican Health and Ageing Study (MHAS), a nationally representative household survey of adults aged 50 years or older (born prior to 1951) living in Mexico. ^13^ MHAS was established to study ageing, health, and socioeconomic determinants of health among older adults in Mexico.^13^

The MHAS baseline survey was conducted in 2001 (Wave 1) using a multistage, stratified sampling design based on the Encuesta Nacional de Empleo (National Employment Survey), with one age-eligible individual randomly selected per household and their spouse included regardless of age. MHAS includes participants from both urban and rural areas across all 32 Mexican states. Individual-level sampling weights are available to account for the complex survey design and to ensure national representativeness.^13^

Follow-up interviews were conducted in 2003 (Wave 2), 2012 (Wave 3), 2015 (Wave 4), and 2018 (Wave 5). Participants were interviewed once per wave, resulting in up to five observations per person over the 17-year follow-up. We defined time as years since the baseline interview, with irregular follow-up intervals corresponding to MHAS waves (0, 2, 11, 14, and 17 years). Health deficits, used to construct the frailty index, and covariates were measured at each wave and reflect participants’ status at the time of interview.

### Frailty Index

To measure frailty, we used a Frailty Index (FI) based on the accumulation of deficits approach.^2^ Following the MHAS standard operating procedure^15^, the FI comprised of 31 equally-weighted health deficits, categorised into seven domains: general health, medically diagnosed conditions, medical symptoms, depressive symptoms, mobility, and limitations in instrumental activities of daily living (IADLs) and activities of daily living (ADLs).

FI deficits were primarily derived from self-reported health measures collected at each survey wave. Most deficits were coded as binary (0= absent, 1 = present); a small number, including self-rated health and self-rated eyesight, were coded on an ordinal scale ranging from 0 to 1 (**Supplementary Table S1**). After dividing the sum of the scores by the number of deficits, FI yields a continuous value bounded between “0” and “1”, with higher values indicating greater frailty.

Frailty was assessed longitudinally, with one FI measurement per participant at each survey wave, resulting in up to five repeated measurements over the 17-year follow-up period. For descriptive analyses, FI was categorised using established thresholds: non-frail (FI ≤ 0.08), pre-frail (FI 0.08-0.24), and frail (FI ≥ 0.25).^16^

To ensure the robustness of FI estimates across waves, two missingness thresholds were applied: i) at the deficit level, variables with ≥5% missingness were excluded from the FI calculation for that wave; and ii) at the observation level, the FI was set to missing for individual participants in a given wave if they had missing data for ≥20% of the included deficits for that wave.

### Covariates

We considered the following covariates as potential confounders: age, household wealth, education, rural residence, smoking status, alcohol use, self-rated health, government health insurance coverage, and cognitive status (**Supplementary Table S2**). Most of covariates were derived from self-reported information collected through interviewer-administered questionnaires, with the exception of cognitive status, which was assessed through performance-based testing.

Baseline age and household wealth were treated as continuous variables and centred at their sample means to improve interpretability and reduce collinearity in interaction models. Household wealth was defined as the total net value of household assets (including housing, financial, and material assets), converted to British Pounds (GBP) to facilitate comparability with international ageing studies, and rescaled so that coefficients represent differences per £1,000.

Education was modelled as a categorical variable (less than upper secondary; upper secondary/vocational; tertiary). Smoking status was categorised as never, former, or current smoker. Rural residence, alcohol consumption (current vs non-drinker), and government health insurance coverage were included as binary variables.

### Statistical analysis

We initially summarised baseline characteristics of the study population and frailty distribution by sex. Continuous variables were summarised using means and standard deviations, and categorical variables using frequencies and percentages.

We modelled longitudinal trajectories of frailty using repeated measures of FI across five waves of the MHAS (full details: **Supplementary Methods**). To characterise longitudinal frailty trajectories and sex differences in frailty accumulation, we fitted survey-weighted linear mixed-effects models with random intercepts and random slopes for time to account for within-participant correlation across repeated measures. The time-scale was time since baseline (2001), with inclusion of quadratic- and cubic-time terms to allow for non-linear frailty trajectories.

Sex and a sex-by-time interaction term were included to assess differences in frailty trajectories between men and women. In this parameterisation, the coefficient for sex represents the mean difference in frailty at baseline (time=0), while the interaction term captures differences in the rate of frailty accumulation over time.

The final longitudinal model included fixed effects for: sex; linear, quadratic, and cubic time; a sex-by-time interaction; baseline age and an age-by-time interaction; household wealth; education; rural residence; physical activity; smoking status; self-rated health; alcohol use and an alcohol-by-time interaction; government health insurance coverage; and cognitive status with a cognitive status-by-time interaction. Covariates were selected *a priori* based on known determinants of frailty and sex differences in those determinants, with the aim of estimating the sex difference in frailty trajectories not explained by between-sex differences in sociodemographic, behavioural and health characteristics.^7^ ^8^ Covariate-by-time interactions were retained where supported by Wald tests (threshold P<0.10), to allow for the possibility that the confounding effect of key covariates on frailty trajectories changed over the follow-up period. Random intercepts and random slopes for time were specified at the participant level, with sex-specific unstructured covariance matrices to allow for heteroskedasticity by sex in the random effects. Alternative random-effects specifications were compared using the Akaike Information Criterion (AIC), with lower values indicating better model fit.^17^ Model fit was assessed using normal quantile plots of residuals and residual-versus-fitted plots, which indicated acceptable fit for the linear mixed-effects model. Estimated frailty trajectories by sex and age group were obtained using post-estimation marginal means from the final longitudinal model.

To investigate sex differences in frailty trajectories in the years preceding death, we fitted survey-weighted linear mixed-effects models restricted to those who died during follow-up (terminal decline model).^6^ ^18^ This analysis mirrors the longitudinal analysis above but with the time scale redefined relative to death rather than study entry: frailty measurements were indexed by years prior to death, with zero representing the year of death and higher values representing a greater number of years before death. Sex and a sex-by-time-to-death interaction were included to assess differences in frailty levels at death and rates of terminal change between men and women. Models included random intercepts and random slopes for time-to-death at the participant level, with sex-specific unstructured covariance matrix. The final terminal decline model included linear, quadratic, and cubic time-to-death terms to allow for non-linear terminal decline and retained the same covariate adjustment structure as the final longitudinal model.

Primary analyses were conducted using multiple imputation with chained equations (40 imputations, stratified by sex) under a missing at random (MAR) assumption. Missing values were imputed for wave-specific frailty indices and covariates (household wealth, education, smoking status, self-rated health, cognitive status, vigorous physical activity, drinking status, and government health insurance). Complete-case analyses were performed as sensitivity analyses to assess the robustness of the findings. All analyses were conducted using Stata version 18.5^19^.

### Secondary analysis

To examine the association between sex and mortality, we fitted survey-weighted Cox proportional hazards models with time since baseline as the timescale, adjusting for baseline frailty index and sociodemographic and health covariates (**Supplementary Methods**).

## RESULTS

A total of 14,154 individuals were interviewed at Wave 1 (2001), of whom 1,669 were excluded as age-ineligible (aged <50 at baseline), leaving 12,485 age-eligible respondents. A further 45 were excluded due to missing health deficits data at all follow-up waves, resulting in a final analytic sample for our study of 12,440 participants.

### Sample Characteristics

The sample included 12,440 adults aged 50-105 years at baseline (mean age, 62.1 years standard deviation [SD] 9.6), including 5,698 men (45.8%) and 6,742 women (54.2%). Most participants lived in urban areas (60.9%). Women were slightly younger than men, had lower educational attainment, and were more likely to have government health insurance coverage (Table 1, **Supplementary Table S3**). Behavioural factors showed marked sex differences: vigorous physical activity was reported by 43.5% of men and 23.2% of women, current smoking by 26.7% of men and 8.7% of women, and alcohol consumption by 46.3% of men and 16.7% of women. Women had higher prevalence of diabetes mellitus (16.9% vs 13.0%) and cognitive impairment (36.9% vs 34.2%).

**Table 1.** Main baseline (Wave 1) characteristics by sex.

| Characteristics | Whole Sample<br>N = 12,440 | Men, n (%)<br>N = 5,698 (45.8) | Women, n (%)<br>N = 6,742 (54.2) |
| --- | --- | --- | --- |
| **Age (years)** |  |  |  |
| Mean (SD) | 62.1 (9.55) | 62.91 (9.6) | 61.96 (9.5) |
| Range (Min, Max) | 50, 105 | 50, 105 | 50, 99 |
| **FI \*** |  |  |  |
| Mean score (SD) | 0.17 (0.12) | 0.16 (0.11) | 0.19 (0.12) |
| **FI categories, n (%) \*** |  |  |  |
| Not frail | 9,546 (75.9) | 4,644 (79.8) | 4,902 (72.6) |
| Frail | 2,566 (20.9) | 860 (16.2) | 1,706 (24.9) |
| **Wealth quintiles, n (%) \*** |  |  |  |
| Low wealth (Q1&Q2) | 4,949 (43.8) | 2,146 (42.4) | 2,803 (45.0) |
| Middle wealth (Q3) | 2,482 (18.2) | 1,182 (19.1) | 1,300 (17.5) |
| High wealth (Q4&Q5) | 4,966 (37.3) | 2,352 (38.0) | 2,614 (36.8) |
| **Government health, n (%) \*** |  |  |  |
| Yes | 7,341 (51.1) | 3,261 (48.8) | 4,080 (52.9) |
| **Rural, n (%)** |  |  |  |
| Yes | 3,188 (39.1) | 1,569 (41.1) | 1,619 (37.4) |
| **Education, n (%) \*** |  |  |  |
| Less than upper secondary | 11,250 (92.2) | 4,926 (89.1) | 6,324 (94.8) |
| Upper secondary and vocational | 265 (1.7) | 195 (3.0) | 70 (0.6) |
| Tertiary | 917 (5.9) | 572 (7.8) | 345 (4.3) |
| **BMI (kg/m<sup>2</sup>), n (%) \*** |  |  |  |
| Underweight ($\leq 18.4$) | 32 (0.3) | 15 (0.3) | 17 (0.3) |
| Normal (18.5-24.9) | 641 (5.8) | 322 (6.5) | 319 (5.2) |
| Pre-obesity (25-29.9) | 941 (8.1) | 483 (8.9) | 458 (7.4) |
| Obesity class I (30-34.9) | 436 (3.2) | 154 (2.0) | 282 (4.3) |
| Obesity class II (35-39.9) | 128 (0.9) | 33 (0.4) | 95 (1.2) |
| Obesity class II ($\geq 40$) | 40 (0.1) | 6 (0.1) | 34 (0.2) |
| **Number of Activities of Daily Living dependencies, n (%) \*** |  |  |  |
| No ADL dependencies (Independent) | 11,034 (90.5) | 5,154 (91.8) | 5,880 (89.3) |
| 1 | 685 (4.3) | 265 (3.5) | 420 (5.0) |
| 2 | 255 (1.6) | 94 (1.5) | 161 (1.6) |
| 3 | 147 (1.0) | 52 (1.0) | 95 (0.9) |
| 4 | 113 (0.8) | 55 (0.6) | 58 (1.1) |
| 5 | 88 (0.7) | 31 (0.3) | 57 (0.9) |
| 6 | 88 (0.6) | 32 (0.6) | 56 (0.7) |
| **Self-report of health, n (%)<sup>\*</sup>** |  |  |  |
| Excellent | 228 (1.7) | 132 (2.3) | 96 (1.2) |
| Very good | 527 (4.3) | 326 (5.5) | 201 (3.4) |
| Good | 3,827 (30.8) | 2,046 (34.7) | 1,781 (27.5) |
| Fair | 5,823 (46.8) | 2,381 (42.3) | 3,442 (50.6) |
| Poor | 2,029 (16.1) | 810 (15.1) | 1,219 (17.1) |
| **Diabetes mellitus, n (%)<sup>\*</sup>** |  |  |  |
| Yes | 1,957 (15.1) | 789 (13.0) | 1,168 (16.9) |
| **Cognitive function status, n (%)<sup>\*</sup>** |  |  |  |
| Normal | 7,085 (52.3) | 3,425 (55.5) | 3,660 (49.5) |
| Normal with Instrumental Impairment | 549 (3.4) | 150 (2.4) | 399 (4.2) |
| Cognitive impairment no dementia (CIND) | 3,867 (35.6) | 1,717 (34.2) | 2,150 (36.8) |
| Dementia | 417 (3.6) | 129 (2.3) | 288 (4.7) |
| **Vigorous activity, n (%)<sup>\*</sup>** |  |  |  |
| Yes | 4,138 (32.5) | 2,431 (43.4) | 1,707 (23.2) |
| **Smoking, n (%)<sup>\*</sup>** |  |  |  |
| Never | 6,929 (57.3) | 1,900 (34.6) | 5,029 (76.6) |
| Former | 3,331 (25.6) | 2,252 (38.5) | 1,079 (14.6) |
| Current Smokers | 2,170 (16.9) | 1,542 (26.7) | 628 (8.7) |
| **Alcohol consumption, n (%)<sup>\*</sup>** |  |  |  |
| Never | 8,536 (69.2) | 2,955 (52.9) | 5,581 (83.1) |
| Ever | 3,848 (30.3) | 2,701 (46.3) | 1,147 (16.7) |

Weighted baseline characteristics results.
n= sample size. SD = standard deviation. FI = Frailty Index BMI = body mass index. GBP, British Pound. Q = quantile.
ADL= activities of daily living, number of reported difficulties with activities of daily living. FI = frailty index.
\* Missing values: FI wave 1 (n=328), Wealth (n=43), Government health (n=29), Education (n=8), BMI (n=10,222), ADL (n=30), SRH (n=6), Diabetes (n=326), Cognitive function (n=522), Vigorous activity (n=101), Smoking (n=10) and Drinking (n=56)

At baseline, mean FI was 0.17 (SD, 0.12), with 76.7% of participants classified as non-frail (FI cut-off <0.25). Women had higher baseline frailty than men (mean FI, 0.19 vs 0.16; mean difference, 0.04; P<0.001) (**Supplementary Table S4, Supplementary Figure S1**).

### Difference in frailty trajectories across sex

Women had higher frailty levels at baseline and accumulated deficits more rapidly over time than men (**Figure 1**). In the fully adjusted mixed-effects model, there was strong evidence that women had higher baseline frailty than men (adjusted mean difference in frailty at baseline β = 0.014 [95% CI, 0.008 to 0.020]; P<0.001). Frailty increased with time in both sexes, with strong evidence of non-linear trajectories (joint test of quadratic and cubic term χ^2^ (2)=88.41; P<0.001). There was strong evidence of a faster rate of frailty accumulation among women than men over follow-up (sex-by-time interaction P=0.008), with the female-male gap in frailty widened over follow-up, increasing from 0.016 FI units at 2 years to 0.029 FI units at 17 years.

**Figure 1.**
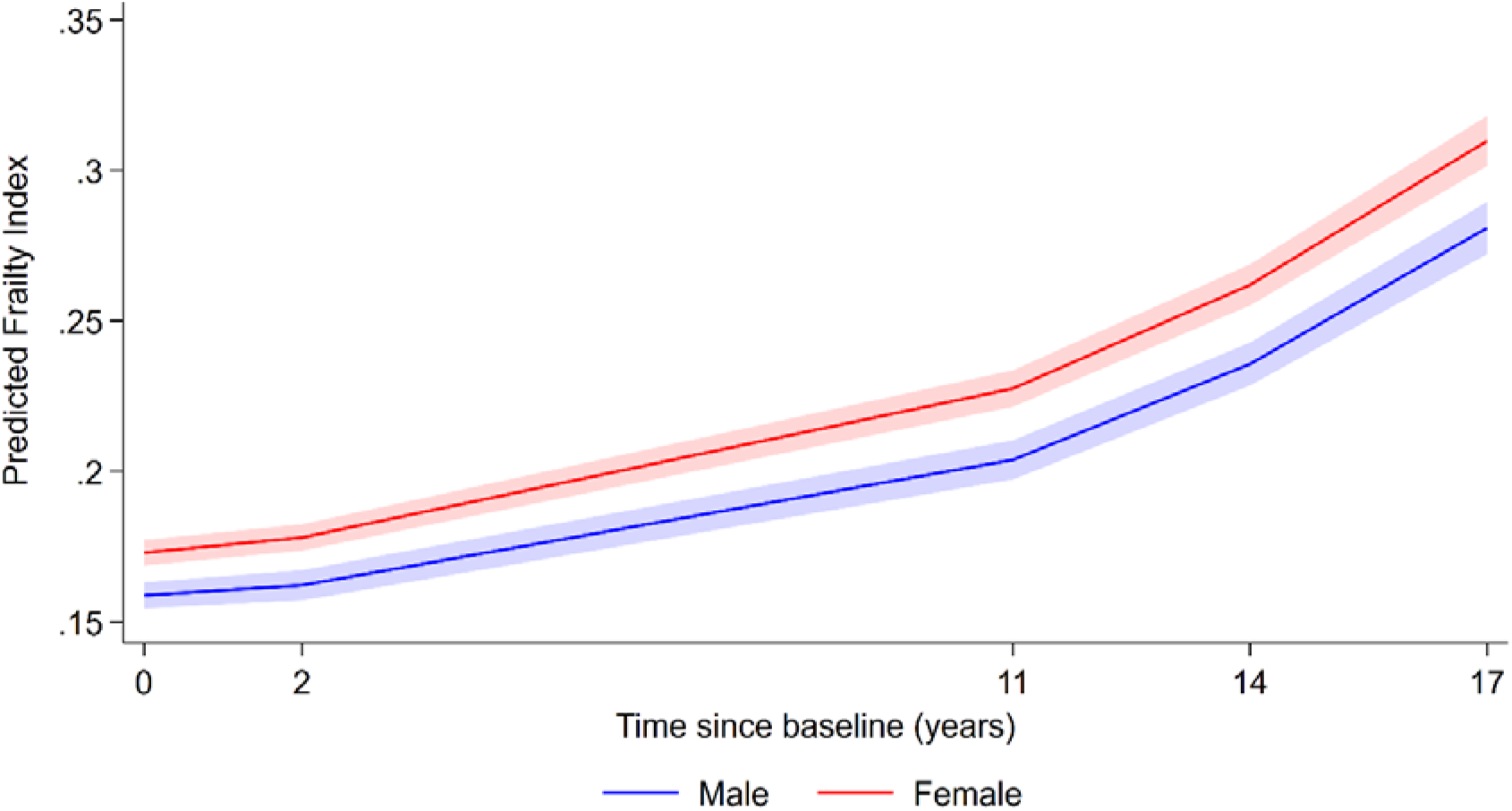
Estimated frailty index trajectories by sex. Solid lines represent model-estimated marginal mean frailty index values, and shaded areas indicate 95% confidence intervals.

Model-estimated marginal mean FI values increased progressively in both sexes; at baseline, mean FI was 0.159 in men and 0.173 in women, rising to 0.281 and 0.310, respectively, at 17 years. At 17 years of follow-up, women had a 27.5% higher predicted frailty index than men, after adjustment for sociodemographic and health covariates.

A sensitivity analysis using a complete-case approach did not materially change the findings (see **Supplementary Table S6**).

### Sex differences in frailty trajectories preceding death (terminal decline)

Among people who died (n = 4,533), frailty trajectories showed marked nonlinearity with accelerating frailty in the years preceding death (**Figure 2**). In the fully adjusted mixed-effects model using multiple imputation for missing covariates and missing frailty values (see methods), women had 0.029 FI points higher adjusted frailty than men at the year of death (95% CI, 0.009 to 0.048; P = 0.004). There was weak evidence of a sex-by-time interaction (β = −0.0016 per year [95% CI, −0.0033 to 0.0001]; P = 0.072), indicating limited evidence that the rate of frailty increases differed between women and men when accounting for missing data.

**Figure 2.**
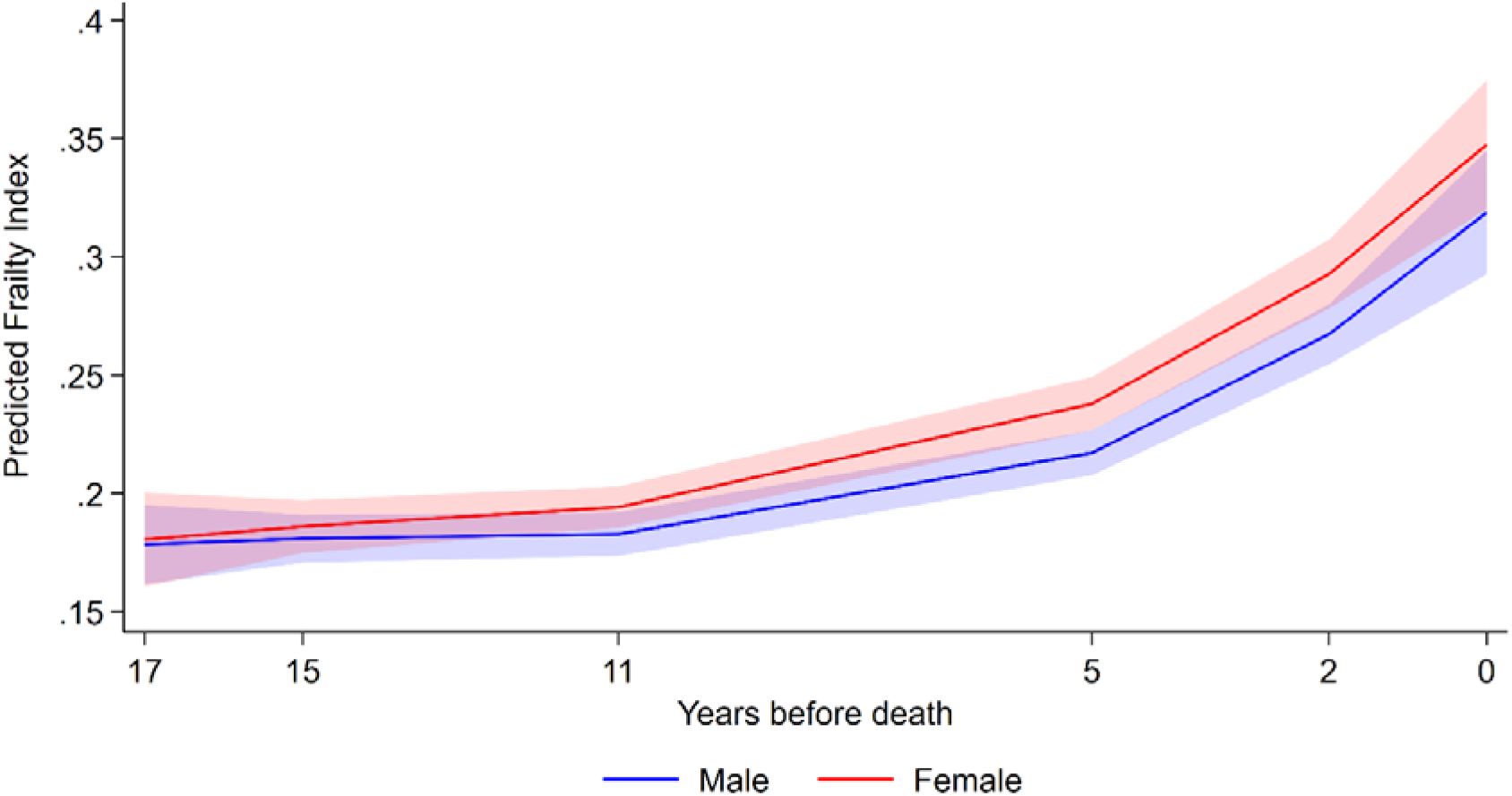
Predicted frailty index trajectories preceding death by sex. Solid lines represent model-estimated marginal mean frailty index values among those who died by years before death (time 0 = death). Shaded areas indicate 95% confidence intervals.

Post-estimation marginal means from the model including the sex-by-time-to death interaction showed that the female-male difference in frailty attenuated with increasing time before death. Women had 0.025 (95% CI, 0.009 to 0.042, P = 0.003) higher frailty index values than men at 2 years before death, 0.021 (95% CI, 0.007 to 0.034, P = 0.003) at 5 years, 0.013 (95% CI, 0.001 to 0.025, P = 0.032) at 10 years, and little difference between the sexes at approximately 15 to 17 years before death.

A sensitivity analysis using a complete-case approach did not materially change the findings (see **Supplementary Table S7**).

### Secondary Analysis

The Cox proportional hazards model showed consistently higher mortality among men compared with women and higher baseline frailty was strongly associated with increased mortality, after adjusting for sociodemographic, behavioural, and health covariates (**Supplementary Figure S2, Table S5**).

## DISCUSSION

In this nationally representative cohort of older adults in Mexico followed over 17 years, we examined sex differences in frailty trajectories and frailty trajectories in the years preceding death. Women had higher frailty at baseline and there was a slowly widening female-male frailty gap across follow-up. Frailty accelerated markedly in the years preceding death; women had higher frailty levels in the years immediately preceding death, with some evidence to suggest a faster rate of increase for women than men. Together, these findings extended deficit accumulation theory to middle-income ageing setting, and provide population-based evidence of sex differences in frailty progression and survival.^2 20^

Our findings are consistent with substantial international evidence demonstrating higher frailty levels among women. Previous population-based studies and meta-analyses have consistently shown that women reach older age with more accumulated health deficits than men.^8 21 22^ This pattern aligns with the male-female health-survival paradox.^21^ Existing explanations include biological differences in immune, hormonal and inflammatory responses, as well as gendered social and behavioural exposures across the life course.^7^ ^10^ ^21^ ^23^ While some cohorts have reported no sex differences in rates of deficit accumulation, others have documented widening frailty gaps with age.^8^ Our findings support a widening sex difference in frailty with age, demonstrating a steeper frailty trajectory among women, extending evidence beyond high-income populations where most prior research has been conducted.

The female survival advantage despite higher frailty burden is consistent with the well-established male-female-survival paradox. These patterns likely reflect interacting biological and social mechanisms operating across the life course, including differences in immune function and hormonal regulation, alongside cumulative social and economic disadvantage.^7^ ^10^ ^23^ ^24^ In LMICs such as Mexico, persistent inequalities in education, employment, and healthcare access may further amplify these processes and contribute to the accumulation of health deficits in women.^24^ ^25^ Emerging clinical evidence also suggests sex-specific differences in frailty-related outcomes, such as sarcopenia, fracture recovery, and heart failure, may also play a role in shaping these patterns.^26 27^

Analyses of sex differences in frailty trajectories preceding death provide further insight into late-life health dynamics between men and women. Frailty trajectories showed marked nonlinearity preceding death, with acceleration in the years immediately preceding death, consistent with terminal decline processes described in ageing research.^6^ ^23^ Women had higher frailty levels in the years immediately preceding death, indicating that they reached death with a greater deficit burden. These findings suggest that women approach death with higher frailty despite their lower mortality hazards.^28^ Such patterns are compatible with theories of sex differences in resilience, disease progression and survival selection.^21 28^ The sex-by-time-to-death interaction did not reach conventional statistical significance, though the coefficient was larger in magnitude than the equivalent interaction in the longitudinal model, suggesting this may reflect lower power in the smaller subsample of decedents rather than a true absence of effect. Considering both chronological and time-to-death analyses together, the female-male frailty gap appears to widen from approximately 10 years before death, suggesting that the divergence in frailty trajectory between men and women extends well beyond the immediate pre-death period.^16^

This study has several strengths. It draws on a large, nationally representative cohort with repeated frailty measurements over nearly two decades, enabling robust modelling of longitudinal trajectories. The use of mixed-effects models allowed estimation of both population-average progression and individual heterogeneity. Incorporating survival analyses and time-to-death modelling provided complementary perspectives on sex differences across ageing and end-of-life processes, reducing bias arising from mortality selection. Survey weighting enhanced population representativeness and supported nationally generalisable estimates. Primary analyses were based on multiple imputation data to address missingness under a missing-at-random assumption.

However, several limitations should be considered. The FI is bounded between 0 and 1 and typically right-skewed; although linear mixed-effects models are widely used and diagnostics indicated acceptable fit, alternative approaches such as beta regression may better capture distributional properties. Deaths were treated as non-informative censoring in primary longitudinal models despite the close relationship between frailty and mortality, which may introduce bias if dropout is informative.

Frailty deficits were derived largely from self-reported measures, and FI construction requires choices regarding deficit selection and thresholds, which may influence comparability. Furthermore, the FI is based on equally weighted deficits, which may not fully reflect the relative contribution of different health conditions to frailty. Frailty, can be measured using different frameworks (e.g., phenotype versus deficit accumulation approaches), and observed sex differences may vary depending on the measurement strategy.^2^ ^29^ ^20^ ^30^ Period effects during follow-up, including healthcare reforms or macroeconomic changes, may also have influenced trajectories and cannot be fully disentangled from ageing processes.^31^ ^32^ Finally, although multiple imputation of missing covariates and missing frailty values supported robustness, the missing-at-random assumption cannot be verified, and residual confounding remains possible.

Nevertheless, these findings have important implications for healthy ageing policy in Mexico and similar middle-income settings. The widening female disadvantage in frailty suggests that current prevention and intervention approaches may not adequately address the structural determinants of health that disproportionately shape frailty risk in women. Structural factors such as educational inequities, caregiving roles and differential healthcare access accumulate across the life course and may contribute to women’s higher frailty burden in later life.^8^ ^12^ ^33^ Effective strategies should, therefore, target upstream social and economic conditions that shape health trajectories, rather than focusing solely on individual-level risk factors. Global frameworks on women, ageing and health explicitly call for policies that improve women’s access to education, decent work, social protection and age friendly health services as a means to-reduce deficit accumulation in later life.^34^ Addressing these requires coordinated action across health, social protection, and economic policy domains.

At the service level, integrated frailty screening within primary care and community health programmes, particularly among women, may facilitate earlier identification of high-risk individuals. Evidence suggests that multi-domain interventions combining exercise, nutrition, medication optimisation and social support can delay onset and progression of frailty, and that tailoring these to sex, for instance prioritising strength, balance and osteoporosis prevention for women, and cardiometabolic risk reduction for men, may enhance effectiveness.^35^ More broadly, reducing rural-urban inequities and promoting education and physical activity may help reduce frailty burden in future cohorts.^1 36^

Future research should investigate mechanisms underlying sex differences in frailty progression (including consideration of sex differences in life-course socioeconomic exposures, infection burden, multimorbidity and healthcare utilisation). Incorporating biomarkers of inflammation, immune function and cardiometabolic health, as well as measures of early-life conditions, would strengthen mechanistic understanding and help to disentangle biological from social pathways.^35^ ^37^ ^38^ Comparative analyses across global cohorts would clarify the generalisability of these findings and identify context-specific drivers of frailty inequalities.

## CONCLUSION

In this nationally representative 17-year longitudinal cohort of older adults in Mexico, sex differences in frailty reflected a widening gap over time. Frailty increased sharply in the years preceding death, with some evidence of a faster rate of increase for women than men. These findings underscore the importance of addressing life-course social and structural determinants of health to reduce sex inequalities in frailty and support equitable healthy ageing in low- and middle-income countries.

## Data Availability

“All data produced in the present study are available online at the Mexican Health and Aging Study (MHAS) website (https://mhasweb.org/), subject to free registration.”

## DECLARATIONS

### Collaborators

Not applicable.

### Contributors

CWG obtained the secondary data used in this study. KA, KEM and CWG conceived of the idea. APB and JN conducted the analysis and created the figures and diagrams. KA derived the frailty index for this study. APB wrote initial draft. All authors (APB, JN, KA, KEM and CWG) contributed to further drafts and approved the final manuscript.

### Disclaimer

The views expressed in this article are the authors’ own and not an official position of their respective institutions.

### Competing

We have no competing interests to declare.

### Provenance and peer review

Not commissioned; externally peer reviewed.

### Data availability statement

Data are available upon request.

### Ethics statements

Patient consent for publication: **Not applicable**.

Ethics approval: Ethics application reference number 32292 /RR/38558

