## Supplemental material for "Sex differences in frailty trajectories among older adults in Mexico: a 17-year longitudinal cohort study"

**Supplementary Materials**

**Sex differences in frailty trajectories and pre-death frailty decline among older adults in Mexico: a 17-year longitudinal cohort study**

**Authors:** Aleixo P. Brunetti^[1]^, Jennifer Nicholas^1^, Kwabena Asare^1^, Kathryn E. Mansfield^[2]^, Charlotte Warren-Gash^1^

### Supplementary Methods

##### Objective 1

We modelled longitudinal trajectories of the frailty index (FI) using repeated measures from five waves of the Mexican Health and Aging Study (MHAS). Survey-weighted linear mixed-effects models (LMMs) with random intercepts and random slopes for time were fitted to account for within-participant correlation across repeated measurements. Sex was included as the primary exposure of interest. The inclusion of a linear, quadratic and cubic terms for time since baseline allowed for a non-linear trajectory in frailty since these higher-order time terms significantly improved model fit versus the model that included only a linear term (χ²(2)=88.41; P<0.001). The model included an interaction between sex and time since baseline, but interactions with the higher-order time terms were not included as sex differences in non-linearity were not supported (χ²(2)=1.53; P=0.465).

The model was adjusted for the following covariates: baseline age, education, household wealth, rural residence, smoking status, alcohol use, self-rated health, government health insurance coverage, and cognitive status. Covariate-by-time interactions were included where necessary, based on evidence from Wald tests (retention threshold P < 0.10). Significant interactions were found for age, drinking status, and cognitive status so these were retained in the model. Other covariate-by-time interactions were not supported (P≥0.10) and so these covariates were included only as main effects.

The final longitudinal model therefore included fixed effects for sex; linear, quadratic, and cubic time; a sex-by-time interaction; baseline age and an age-by-time interaction; household wealth; education; rural residence; physical activity; smoking status; self-rated health; alcohol use and an alcohol-by-time interaction; government health insurance coverage; and cognitive status with a cognitive status-by-time interaction.

Models included random intercepts and random slopes for time at the participant level. To select the random-effects covariance structures, these were compared using Akaike Information Criterion (AIC) from the survey-weighted pseudo-likelihood. Structures requiring equal time spacing (e.g., autoregressive, Toeplitz) were not considered due to irregular follow-up intervals, and identity and exchangeable structures were deemed inappropriate for intercept-slope pairs. An unstructured covariance structure provided the best fit (AIC = −81,256,652 vs −81,057,550 for independent). Heteroskedasticity by sex was evaluated at both the residual (level 1) and random-effects (level 2) levels. At level 1, when residual variance was allowed to differ by sex, which modestly improved model fit (AIC = −81,260,375). At level 2, the inclusion of sex-specific covariance matrices for men and women produced a better fit (AIC = −81,372,516) and was retained in the final longitudinal model as the random-effects structure. Therefore, the final model included random intercepts and random slopes for time at the participant level, with sex-specific unstructured covariance matrices to allow for level 2 heteroskedasticity.

Model assumptions were assessed using residual and random-effects diagnostics, including normal distribution quantile-quantile plots and residual-versus-fitted plots, which indicated some remaining heteroskedasticity. Robust standard errors, clustered at the participant level, were used to account for remaining heteroskedasticity.

From the final longitudinal model, predicted marginal mean FI values were obtained by sex and time since baseline using post-estimation marginal means. Differences in FI between women and men at selected time points were calculated from these marginal predictions, with corresponding 95% confidence intervals derived from the model-based variance estimates.

##### Objective 2

To examine sex differences in frailty trajectories preceding death, analyses were restricted to participants who died during follow-up. The longitudinal dataset was restructured using a backward-time-to-death scale, defined as the number of years between each frailty assessment and the date of death, with time-to-death coded as 0 at death and increasing values representing earlier time points.

Linear mixed-effects models analogous to those used in Objective 1 were fitted, with the FI as the outcome and linear, quadratic, and cubic time-to-death terms to capture non-linear terminal decline trajectories. Sex and a sex-by-time-to-death interaction were included to assess sex differences in terminal frailty accumulation. Models were adjusted for the same baseline socioeconomic, behavioural, and health-related covariates as in Objective 1 (baseline age, education, household wealth, rural residence, smoking status, alcohol use, self-rated health, government health insurance coverage, and cognitive status). In addition, consistent with final longitudinal model, interactions of age, alcohol use, and cognitive status with time-to-death were included.

Random intercepts and random slopes for time-to-death were specified at the participant level, with sex-specific unstructured covariance matrices to allow heterogeneity in frailty levels at death and terminal slopes. Survey calibration weights were applied, and robust standard errors were used to account for clustering and heteroskedasticity. Model-based predicted frailty trajectories by sex were estimated using predictive margins at selected years before death (0, 2, 5, 10, 15, and 17 years).

#### Handling missing data

Multiple imputation by chained equations was conducted under a missing-at-random assumption. 40 imputed datasets were generated after 20 burn-in iterations, incorporating survey calibration weights. Predictive mean matching with 10 nearest neighbours was used for continuous variables (wave 1 frailty index [w1fi], w2fi, w3fi, w4fi, w5fi, and household wealth in GBP), multinomial logistic regression for nominal categorical variables (smoke status and education), ordinal logistic regression for ordered categorical variables (self-rated health and cognitive status), and logistic regression for binary variables (vigorous physical activity, drinking status, and health insurance). Frailty was imputed at the wave level (w1fi-w5fi) and then used to construct the longitudinal FI outcome (wfi) in the long-format analysis dataset. The fully observed variables included in the imputation model were sex (used for stratification), baseline age (age), and rural residence (rural), and survey calibration weights were incorporated using probability weights (w1respwt). Imputation was stratified by sex to allow the model to be compatible with the presence of sex specific frailty trajectories and effects of the imputed variables on these trajectories. The same imputation procedure was used for the time-to-death analysis. Frailty indices were first imputed in wide format (w1fi-w5fi), after which time-to-death variables were derived and the dataset reshaped to long format for the terminal decline models among decedents.

The primary longitudinal mixed-effects model with survey weights and robust standard errors was refitted to each imputed dataset and results combined using Rubin’s rules.

#### Secondary Analysis

To examine the association between sex and mortality, we fitted survey-weighted Cox proportional hazards models with time since baseline as the timescale. Sex was the primary exposure, baseline frailty index (centered) was included as a covariate to assess whether the female survival advantage persisted after accounting for women's higher frailty burden. A sex-by-frailty interaction term was added to assess effect modification but was not statistically supported and was not retained in the final model. Models were additionally adjusted for age, household wealth, education, rural residence, vigorous physical activity, smoking status, self-rated health, drinking status, government health insurance coverage, and cognitive status.

The proportional hazards assumption was assessed using global and covariate-specific tests based on scaled Schoenfeld residuals and graphical inspection of Schoenfeld and log-log survival plots. Potential non-linearity in the functional form of continuous covariates, including frailty, was evaluated using Martingale residuals and smoothed plots. No strong evidence of non-linearity was observed, and continuous covariates were retained in linear form in the final model. Time-varying effects of frailty and other covariates were explored using models with time-dependent coefficients; no meaningful departures from proportional hazards were detected. Robust variance estimation accounting for clustering at the individual level was used throughout.

**Sensitivity analysis**

A complete-case analysis was conducted as a sensitivity analysis to assess the robustness of the primary multiple imputation findings.

#### Analytical approach

All analyses were conducted using a pre-specified and reproducible analytical workflow in Stata version 18.5. Data management, variable derivation, modelling, and figure generation were performed using scripted do-files to ensure consistency and reproducibility. Longitudinal and survival datasets were structured according to the MHAS wave design, and identical modelling procedures were applied across complete-case and imputed datasets. Results are reported in accordance with STROBE recommendations for longitudinal cohort studies.

### Supplementary Table S1: Health deficits, coding and wave availability used to construct the frailty in the MHAS

|  | **Deficit** | **Cut-off** | **Deficit availability (> 5% non-missing)** |
| --- | --- | --- | --- |
| **General health** | | | |
| 1 | R Self-report of health | Excellent = 0, Very good = 0.25, Good = 0.5, Fair = 0.75, Poor = 1 | Waves 1-5 |
| **Medically diagnosed conditions** | | | |
| 2 | R Ever had high blood pressure | No = 0, Yes = 1 | Waves 1-5 |
| 3 | R Ever had diabetes | No = 0, Yes = 1 | Waves 1-5 |
| 4 | R Ever had cancer | No = 0, Yes = 1 | Waves 1-5 |
| 5 | R Ever had respiratory disease, incl. asthma | No = 0, Yes = 1 | Waves 1-5 |
| 6 | R Ever had heart attack | No = 0, Yes = 1 | Waves 1-5 |
| 7 | R Ever had stroke | No = 0, Yes = 1 | Waves 1-5 |
| 8 | R Ever had arthritis | No = 0, Yes = 1 | Waves 1-5 |
| 9 | R Ever fractured a bone (including hip) since age 50 | No = 0, Yes = 1 | Waves 1-5 |
| **Medical symptoms** | | | |
| 10 | R Fallen down last 2 years | No = 0, Yes = 1 | Waves 1-5 |
| 11 | R Self-rated eyesight | Excellent = 0, Very good = 0.2, Good = 0.4, Fair = 0.6, Poor = 0.8, Legally Blind = 1 | Waves 1-5 |
| 12 | R Self-rated hearing | Excellent = 0, Very good = 0.2, Good = 0.4, Fair = 0.6, Poor = 0.8, Legally Deaf = 1 | Waves 1, 2, 4, 5 |
| 13 | R Severe fatigue | No = 0, Yes = 1 | Waves 1-5 |
| 14 | R Difficulty breathing | No = 0, Yes = 1 | Waves 1-5 |
| 15 | R Pain in lower limbs while (or after) walking | No = 0, Yes = 1 | Waves 1-2 |
| 16 | R Stomach pain, indigestion, diarrhoea | No = 0, Yes = 1 | Waves 1-5 |
| 17 | R Frequent problems with pain | No = 0, Yes = 1 | Waves 1-5 |
| 18 | R Leaks urine when coughing (last 2 yrs) | No = 0, Yes = 1 | Waves 1-5 |
| **Depressive symptoms (CES-D)** | | | |
| 19 | Felt depressed | No = 0, Yes = 1/9 | Waves 1-5 |
| 20 | Everything an effort | No = 0, Yes = 1/9 | Waves 1-5 |
| 21 | Sleep was restless | No = 0, Yes = 1/9 | Waves 1-5 |
| 22 | Felt happy | No = 1/9, Yes = 0 | Waves 1-5 |
| 23 | Felt lonely | No = 0, Yes = 1/9 | Waves 1-5 |
| 24 | Felt sad | No = 0, Yes = 1/9 | Waves 1-5 |
| 25 | Enjoyed life | No = 1/9, Yes = 0 | Waves 1-5 |
| 26 | Felt tired | No = 0, Yes = 1/9 | Waves 1-5 |
| 27 | Had a lot of energy | No = 1/9, Yes = 0 | Waves 1-5 |
| **Mobility** | | | |
| 28 | R Difficulty – Picking up a coin | No = 0, Yes = 1 | Waves 1-5 |
| 29 | R Difficulty – Dressing | No = 0, Yes = 1 | Waves 1-5 |
| 30 | R Difficulty – Walking several blocks | No = 0, Yes = 1 | Waves 1-5 |
| 31 | R Difficulty – Walking across room | No = 0, Yes = 1 | Waves 1-5 |
| **Activities of Daily Living (ADL)** | | | |
| 32 | R Difficulty – Bathing or showering | No = 0, Yes = 1 | Waves 1-5 |
| 33 | R Difficulty – Eating | No = 0, Yes = 1 | Waves 1-5 |
| 34 | R Difficulty – Getting in/out of bed | No = 0, Yes = 1 | Waves 1-5 |
| 35 | R Difficulty – Using the toilet | No = 0, Yes = 1 | Waves 1-5 |
| **Instrumental Activities of Daily Living (IADL)** | | | |
| 36 | R Difficulty – Preparing hot meals | No = 0, Yes = 1 | Waves 1-5 |
| 37 | R Difficulty – Shopping for groceries | No = 0, Yes = 1 | Waves 1-5 |
| 38 | R Difficulty – Taking medications | No = 0, Yes = 1 | Waves 1-5 |
| 39 | R Difficulty – Managing money | No = 0, Yes = 1 | Waves 1-5 |
| CES-D = Center for Epidemiologic Studies Depression Scale. R = respondent.  A total of 39 health variables were used to construct the frailty index. 9 CES-D depressive symptom items were coded as 1/9 and combined into a single depressive symptoms deficit, resulting in a maximum of 31 deficits.  Due to variable availability across waves, the number of available variables was 39 in waves 1-2, 37 in wave 3, and 38 in waves 4-5.  The frailty index (FI) was calculated as:  FI = sum of deficits present / number of available deficits for that observation in that wave. | | | |

### Supplementary Table S2: Variables of interest in the MHAS study

| **Variable** | **Type** | **Categories/ Range** | **Measurement Details** |
| --- | --- | --- | --- |
| Age | Continuous | Years | Age at baseline |
| Month and Year of Birth | Continuous |  | Recorded in months and years |
| Month and Year of Death | Continuous |  | Recorded in months and years |
| Person-level analysis weight | Continuous |  | Survey calibration weight at baseline |
| Time | Continuous | 0, 2, 11, 14, 17 years | Years elapsed since 2001 baseline for each wave. |
| Biological Sex | Binary | 1 = Male, 2 = Female |  |
| Education | Categorical | 1= Less than upper secondary, 2 = Upper secondary and Vocational, 3= Tertiary | Highest level attained |
| Place of residence | Binary | 0 = Urban, 1 = Rural | Based on location of household |
| Household Wealth Index | Continuous | GBP (in 1,000s) | Total household wealth at baseline. |
| Vigorous Physical Activity | Binary | 0 = No, 1 = Yes | ≥3 times per week |
| Currently Drink | Binary | 0 = No, 1 = Yes | Alcohol use at time of survey |
| Ever Drink | Binary | 0 = No, 1 = Yes | Lifetime alcohol use |
| Alcohol Consumption per Week | Continuous | 0–7 days | Frequency of alcohol use |
| Ever Smoke | Binary | 0 = No, 1 = Yes | Lifetime smoking status |
| Current Smoke | Binary | 0 = No, 1 = Yes | Smoking at time of survey |
| Activities of Daily Living | Continuous | 1 to 6 difficulties | Number of reported difficulties |
| Self-Rated Health | Categorical | 1 = Excellent, 2 = Very good, 3 = Good, 4 = Fair, 5 = Poor | Self-reported health status |
| Government Health Insurance | Binary | 0 = No, 1 = Yes | Any government program coverage |
| Cognitive Status | Categorical | 1 = Normal, 2 = Normal + instrumental impairment, 3 = Cognitive impairment (no dementia), 4 = Dementia | Based on cognitive assessment |

### Supplementary Table S3: Weighted baseline characteristics of the study population by sex

|  | **Whole Sample** | **Men, n (%)** | **Women, n (%)** |  |
| --- | --- | --- | --- | --- |
| **Characteristics** | **N = 12,440** | **N = 5,698 (45.8)** | **N= 6,742 (54.2)** | **p-value** |
| **Age (years)** |  |  |  | <0.003 |
| Mean (SD) | 62.1 (9.55) | 62.91 (9.6) | 61.96 (9.5) |  |
| Range (Min, Max) | 50, 105 | 50, 105 | 50, 99 |  |
| **FI Wave 1** |  |  |  | < 0.0001 |
| Score (SD) | 0.17 (0.12) | 0.16 (0.11) | 0.19 (0.12) |  |
| Missing, n (%) | 328 (2.64) | 194 (3.4) | 134 (2.0) |  |
| **FI categories Wave 1, n (%)** | |  |  | < 0.0001 |
| Not frail | 9,546 (75.94) | 4,644 (79.84) | 4,902 (72.63) |  |
| Frail | 2,566 (20.95) | 860 (16.28) | 1,706 (24.91) |  |
| Missing | 328 (3.11) | 194 (3.87) | 134 (2.46) |  |
| **Wealth quintiles, n (%)** | |  |  | < 0.3121 |
| Low wealth (Q1&Q2) | 4,949 (43.86) | 2,146 (42.46) | 2,803 (45.04) |  |
| Middle wealth (Q3) | 2,482 (18.27) | 1,182 (19.12) | 1,300 (17.54) |  |
| High wealth (Q4&Q5) | 4,966 (37.35) | 2,352 (38.01) | 2,614 (36.80) |  |
| Missing | 43 (0.52) | 18 (0.41) | 25 (0.62) |  |
| **Government health, n (%)** | |  |  | < 0.0216 |
| No | 5,070 (48.73) | 2,425 (50.95) | 2,645 (46.84) |  |
| Yes | 7,341 (51.09) | 3,261 (48.86) | 4,080 (52.99) |  |
| Missing | 29 (0.18) | 12 (0.19) | 17 (0.17) |  |
| **Rural, n (%)** |  |  |  | < 0.0262 |
| No | 9,252 (60.90) | 4,129 (58.90) | 5,123 (62.60) |  |
| Yes | 3,188 (39.10) | 1,569 (41.10) | 1,619 (37.40) |  |
| **Education, n (%)** |  |  |  | < 0.0001 |
| Less than upper secondary | 11,250 (92.23) | 4,926 (89.12) | 6,324 (94.87) |  |
| Upper secondary and vocational | 265 (1.76) | 195 (3.01) | 70 (0.69) |  |
| Tertiary | 917 (5.94) | 572 (7.83) | 345 (4.33) |  |
| Missing | 8 (0.08) | 5 (0.03) | 3 (0.11) |  |
| **BMI (Kg/m^2^), n (%)** |  |  |  | < 0.0001 |
| Underweight (< 18) | 32 (0.35) | 15 (0.38) | 17 (0.31) |  |
| Normal (18.5-24.9) | 641 (5.84) | 322 (6.52) | 319 (5.27) |  |
| Pre-obesity (25-29) | 941 (8.14) | 483 (8.91) | 458 (7.49) |  |
| Obesity class I (30–34.9) | 436 (3.28) | 154 (2.03) | 282 (4.35) |  |
| Obesity class II (35–39.9) | 128 (0.90) | 33 (0.46) | 95 (1.28) |  |
| Obesity class II (≥ 40) | 40 (0.16) | 6 (0.07) | 34 (0.24) |  |
| Missing, n (%) | 10,222 (81.3) | 4,685 (81.63) | 5,537 (81.06) |  |
| **ADL, n (%)** |  |  |  | < 0.0383 |
| No ADLs (Independent) | 11,034 (90.53) | 5,154 (91.88) | 5,880 (89.39) |  |
| 1 | 685 (4.36) | 265 (3.58) | 420 (5.03) |  |
| 2 | 255 (1.63) | 94 (1.58) | 161 (1.67) |  |
| 3 | 147 (1.02) | 52 (1.06) | 95 (0.98) |  |
| 4 | 113 (0.89) | 55 (0.64) | 58 (1.11) |  |
| 5 | 88 (0.70) | 31 (0.39) | 57 (0.95) |  |
| 6 | 88 (0.69) | 32 (0.67) | 56 (0.70) |  |
| Missing | 30 (0.18) | 15 (0.21) | 15 (0.15) |  |
| **SRH, n (%)** |  |  |  | < 0.0001 |
| Excellent | 228 (1.72) | 132 (2.32) | 96 (1.21) |  |
| Very good | 527 (4.38) | 326 (5.52) | 201 (3.41) |  |
| Good | 3,827 (30.84) | 2,046 (34.70) | 1,781 (27.57) |  |
| Fair | 5,823 (46.85) | 2,381 (42.34) | 3,442 (50.69) |  |
| Poor | 2,029 (16.15) | 810 (15.07) | 1,219 (17.07) |  |
| Missing | 6 (0.05) | 3 (0.05) | 3 (0.05) |  |
| **Diabetes, n (%)** |  |  |  | < 0.0004 |
| No | 10,157 (81.64) | 4,718 (82.90) | 5,439 (80.57) |  |
| Yes | 1,957 (15.12) | 789 (13.02) | 1,168 (16.91) |  |
| Missing | 326 (3.24) | 191 (4.09) | 135 (2.52) |  |
| **Cognitive Function Status, n (%)** | |  |  | < 0.0001 |
| Normal | 7,085 (52.31) | 3,425 (55.55) | 3,660 (49.56) |  |
| Normal with Instrumental Impairment | 549 (3.42) | 150 (2.43) | 399 (4.26) |  |
| CIND | 3,867 (35.67) | 1,717 (34.23) | 2,150 (36.89) |  |
| Dementia | 417 (3.65) | 129 (2.31) | 288 (4.79) |  |
| Missing | 522 (4.96) | 277 (5.48) | 245 (4.51) |  |
| **Vigorous activity, n (%)** |  |  |  | < 0.0001 |
| No | 8,201 (66.79) | 3,217 (55.77) | 4,984 (76.14) |  |
| Yes | 4,138 (32.51) | 2,431 (43.45) | 1,707 (23.22) |  |
| Missing | 101 (0.71) | 50 (0.78) | 51 (0.64) |  |
| **Smoking, n (%)** |  |  |  | < 0.0001 |
| Never | 6,929 (57.34) | 1,900 (34.64) | 5,029 (76.61) |  |
| Former | 3,331 (25.61) | 2,252 (38.55) | 1,079 (14.61) |  |
| Current Smokers | 2,170 (16.98) | 1,542 (26.71) | 628 (8.71) |  |
| Missing | 10 (0.08) | 4 (0.10) | 6 (0.07) |  |
| **Drinking, n (%)** |  |  |  | < 0.0001 |
| Never | 8,536 (69.27) | 2,955 (52.90) | 5,581 (83.17) |  |
| Ever Drink | 3,848 (30.31) | 2,701 (46.31) | 1,147 (16.72) |  |
| Missing | 56 (0.42) | 42 (0.79) | 14 (0.11) |  |
| Weighted baseline characteristics results. n= sample size. SD = standard deviation. FI = Frailty Index BMI = body mass index. GBP, British Pound. Q = quantile. ADL= Activities of Daily Living. SRH = self-report of health. FI = frailty index. CIND= Cognitive impairment no dementia. | | | | |

### Supplementary Table S4: Frailty index and frailty status by sex across Waves 1-5.

| **3.1: Male participants** | | | | | |
| --- | --- | --- | --- | --- | --- |
|  | **Wave 1** | **Wave 2** | **Wave 3** | **Wave 4** | **Wave 5** |
| **Characteristics** | **N=5,698** | **N=5,698** | **N=5,698** | **N=5,698** | **N=5,698** |
| **Frailty Index** |  |  |  |  |  |
| Mean (SD) | 0.15 (0.11) | 0.16 (0.11) | 0.17 (0.12) | 0.20 (0.12) | 0.22 (0.12) |
| Median (IQR) | 0.12 (0.08,0.20) | 0.13 (0.08,0.22) | 0.15 (0.09,0.23) | 0.17 (0.11,0.26) | 0.20 (0.13,0.28) |
| Range | 0.00, 0.82 | 0.00, 0.81 | 0.00, 0.88 | 0.00, 0.79 | 0.03, 0.79 |
| Missing, n (%) | 194 (3.4) | 1081 (19.0) | 2573 (45.2) | 2839 (49.8) | 3474 (61.0) |
| **Frailty Category** | |  |  |  |  |
| Not Frail | 4644 (79.8) | 3797 (66.6) | 2464 (43.2) | 2052 (36.0) | 1488 (26.1) |
| Frail | 860 (16.3) | 820 (14.4) | 661 (11.6) | 807 (14.2) | 736 (12.9) |
| Missing, n (%) | 194 (3.9) | 1081 (19.0) | 2573 (45.2) | 2839 (49.8) | 3474 (61.0) |
| **3.2: Female participants** | | | | | |
|  | **Wave 1** | **Wave 2** | **Wave 3** | **Wave 4** | **Wave 5** |
| **Characteristics** | **N=6,742** | **N=6,742** | **N=6,742** | **N=6,742** | **N=6,742** |
| **Frailty Index** | |  |  |  |  |
| Mean (SD) | 0.19 (0.12) | 0.20 (0.12) | 0.22 (0.13) | 0.25 (0.14) | 0.27 (0.14) |
| Median (IQR) | 0.16 (0.10,0.25) | 0.18 (0.11,0.26) | 0.19 (0.12,0.29) | 0.23 (0.15,0.33) | 0.25 (0.17,0.34) |
| Range | 0.00, 0.81 | 0.00, 0.81 | 0.00, 0.88 | 0.01, 0.84 | 0.03, 0.87 |
| Missing, n (%) | 134 (2.0) | 925 (13.7) | 2680 (39.8) | 3042 (45.1) | 3826 (56.7) |
| **Frailty Category** | |  |  |  |  |
| Not Frail | 4902 (72.6) | 4216 (62.5) | 2681 (39.8) | 2101 (31.2) | 1434 (21.3) |
| Frail | 1706 (24.9) | 1601 (23.7) | 1381 (20.5) | 1599 (23.7) | 1482 (22.0) |
| Missing, n (%) | 134 (2.5) | 925 (13.7) | 2680 (39.8) | 3042 (45.1) | 3826 (56.7) |
| Weighted results | | | | | |
| SD = standard deviation. N = sample size. IQR = interquartile range | | | | | |

### Supplementary Figure S1: Distribution of the Frailty Index across MHAS Waves 1 to 5 stratified by sex

| 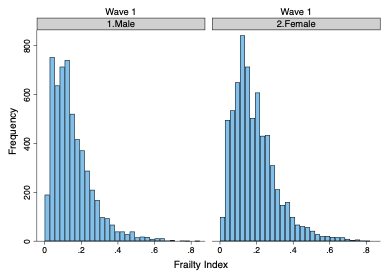 | 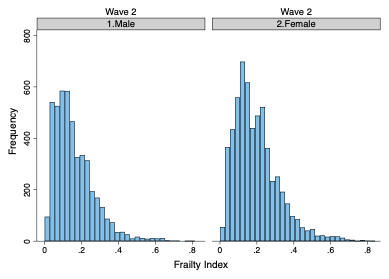 |
| --- | --- |
| 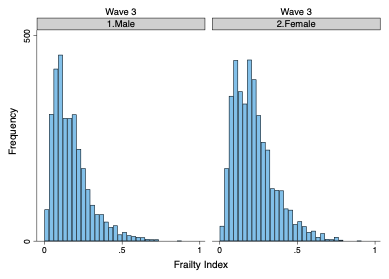 | 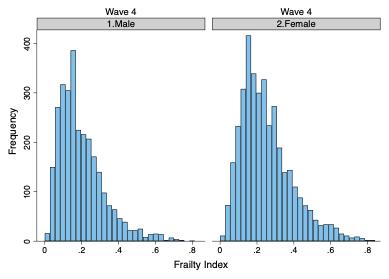 |
| 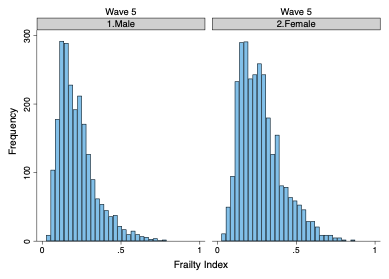 | |

### Supplementary Figure S2: Adjusted survival curves by sex from the Cox proportional hazards model

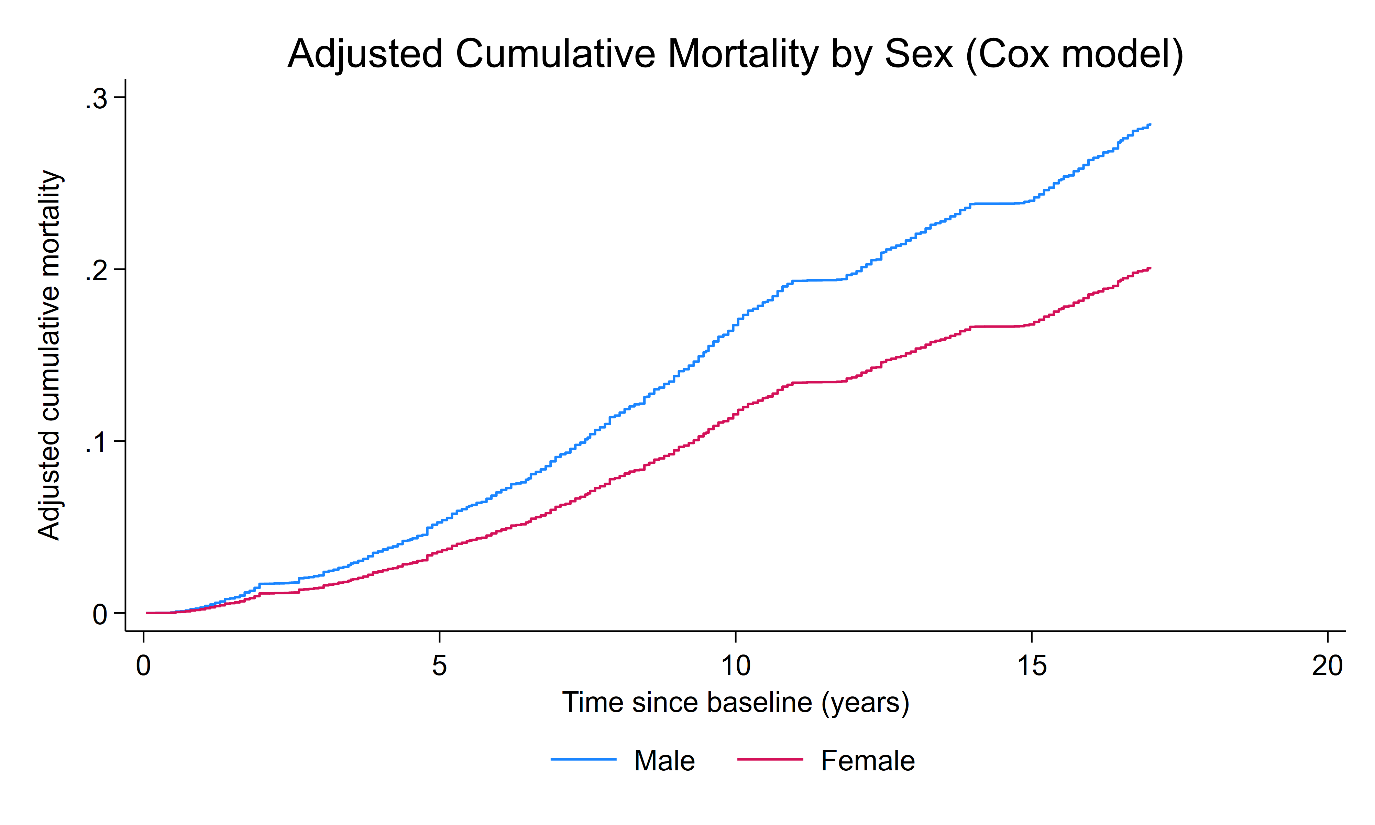

### Supplementary Table S5: Survey-weighted Cox regression HR (95%Cis) for mortality

| **Predictor (units; base)** | **HR** | **SE** | **95% CI** | **p-value** |
| --- | --- | --- | --- | --- |
| **Sex** |  |  |  |  |
| └ Male (base) | 1 (ref.) | — | — | — |
| └ Female | 0.6702 | 0.0556 | 0.5696, 0.7886 | <0.001 |
| **Frailty index (per 0.10 increase)** | 1.3977 | 0.0364 | 1.3282, 1.4709 | <0.001 |
| **Age (centred)** | 1.0776 | 0.0042 | 1.0694, 1.0858 | <0.001 |
| **Wealth (GBP; centred)** | 0.999999 | 0.000002 | 0.999995, 1.000002 | 0.531 |
| **Education** |  |  |  |  |
| └ Less than upper secondary (base) | 1 (ref.) | — | — | — |
| └ Upper secondary and vocational | 0.7929 | 0.2105 | 0.4712, 1.3343 | 0.382 |
| └ Tertiary | 1.2817 | 0.292 | 0.8200, 2.0032 | 0.276 |
| **Rural residence** |  |  |  |  |
| └ No (base) | 1 (ref.) | — | — | — |
| └ Yes | 0.9743 | 0.0776 | 0.8335, 1.1388 | 0.743 |
| **Vigorous activity** |  |  |  |  |
| └ No (base) | 1 (ref.) | — | — | — |
| └ Yes | 0.876 | 0.0702 | 0.7488, 1.0249 | 0.098 |
| **Smoking status** |  |  |  |  |
| └ Never (base) | 1 (ref.) | — | — | — |
| └ Former | 1.1492 | 0.0976 | 0.9730, 1.3572 | 0.102 |
| └ Current | 1.2943 | 0.1309 | 1.0616, 1.5780 | 0.011 |
| **Self-rated health** |  |  |  |  |
| └ Excellent (base) | 1 (ref.) | — | — | — |
| └ Very good | 1.3566 | 0.6124 | 0.5601, 3.2862 | 0.499 |
| └ Good | 0.9891 | 0.3413 | 0.5029, 1.9454 | 0.975 |
| └ Fair | 0.9023 | 0.3111 | 0.4590, 1.7734 | 0.765 |
| └ Poor | 1.0521 | 0.3712 | 0.5269, 2.1008 | 0.886 |
| **Alcohol consumption** |  |  |  |  |
| └ Never (base) | 1 (ref.) | — | — | — |
| └ Current | 1.0492 | 0.0929 | 0.8820, 1.2481 | 0.587 |
| **Health insurance (higov)** |  |  |  |  |
| └ No (base) | 1 (ref.) | — | — | — |
| └ Yes | 1.1363 | 0.0916 | 0.9702, 1.3308 | 0.113 |
| **Cognitive status** |  |  |  |  |
| └ Normal (base) | 1 (ref.) | — | — | — |
| └ Normal with instrumental impairment | 0.9889 | 0.136 | 0.7553, 1.2948 | 0.935 |
| └ CIND | 1.115 | 0.0876 | 0.9558, 1.3007 | 0.166 |
| └ Dementia | 1.6953 | 0.229 | 1.3009, 2.2093 | <0.001 |
| Hazard ratios (HRs) are derived from a survey-weighted Cox proportional hazards regression model. The frailty index is scaled per 0.10 increase. Age and wealth are centred at the sample mean. The reference group is male, less than upper secondary education, non-rural residence, no vigorous activity, never smoker, excellent self-rated health, never drinker, no government health insurance, and normal cognitive status. SE = linearized standard error. CI = confidence interval. | | | | |

### Supplementary Table S6: Longitudinal Mixed-effects estimates of frailty trajectories comparing multiple imputation and complete-case analysis.

|  | **Longitudinal Model: Multiple imputation** | | | | **Longitudinal Model: Complete-case analysis** | | | |
| --- | --- | --- | --- | --- | --- | --- | --- | --- |
| **Predictor (units; base)** | **Coef.** | **SE** | **95% CI** | **p-value** | **Coef.** | **SE** | **95% CI** | **p-value** |
| **Fixed effects** |  |  |  |  |  |  |  |  |
| **Sex** |  |  |  |  |  |  |  |  |
| └ Male (base) | 0 (ref.) | — | — | — | 0 (ref.) | — | — | — |
| └ Female | 0.01412 | 0.00312 | 0.00800, 0.02025 | <0.001 | 0.01506 | 0.00319 | 0.00881, 0.02131 | <0.001 |
| **Time (years since baseline)** | 0.00227 | 0.00127 | −0.00022, 0.00476 | 0.074 | 0.0026 | 0.0013 | 0.00005, 0.00516 | 0.046 |
| **Time²** | 0.00006 | 0.00017 | −0.00028, 0.00040 | 0.735 | 0.00001 | 0.00018 | −0.00034, 0.00037 | 0.938 |
| **Time³** | 0.0000163 | 0.0000064 | 0.0000037, 0.0000289 | 0.011 | 0.000018 | 0.0000066000 | 0.0000051, 0.0000309 | 0.006 |
| **Female × Time** | 0.00087 | 0.00033 | 0.00022, 0.00152 | 0.008 | 0.00088 | 0.00034 | 0.00022, 0.00154 | 0.009 |
| **Age (centred)** | 0.00131 | 0.00017 | 0.00098, 0.00165 | <0.001 | 0.00108 | 0.00017 | 0.00074, 0.00142 | <0.001 |
| **Age × Time** | 0.00029 | 0.00003 | 0.00023, 0.00034 | <0.001 | 0.00029 | 0.00003 | 0.00024, 0.00035 | <0.001 |
| **Wealth (GBP; centred)** | −0.0000000260 | 0.0000000596 | −0.0000001430, 0.0000000907 | 0.662 | −0.0000000181 | 0.000000061 | −0.0000001380, 0.0000001010 | 0.767 |
| **Education** |  |  |  |  |  |  |  |  |
| └ < Upper secondary (base) | 0 (ref.) | — | — | — | 0 (ref.) | — | — | — |
| └ Upper secondary/vocational | −0.01178 | 0.00813 | −0.02772, 0.00416 | 0.147 | −0.01200 | 0.00823 | −0.02814, 0.00414 | 0.145 |
| └ Tertiary | −0.01474 | 0.00428 | −0.02313, −0.00635 | 0.001 | −0.01382 | 0.00428 | −0.02221, −0.00543 | 0.001 |
| **Rural residence** |  |  |  |  |  |  |  |  |
| └ No (base) | 0 (ref.) | — | — | — | 0 (ref.) | — | — | — |
| └ Yes | 0.00667 | 0.003 | 0.00079, 0.01256 | 0.026 | 0.00706 | 0.00307 | 0.00104, 0.01309 | 0.022 |
| **Vigorous activity** |  |  |  |  |  |  |  |  |
| └ No (base) | 0 (ref.) | — | — | — | 0 (ref.) | — | — | — |
| └ Yes | −0.00500 | 0.00275 | −0.01039, 0.00039 | 0.069 | −0.00439 | 0.0028 | −0.00987, 0.00110 | 0.117 |
| **Smoking status** |  |  |  |  |  |  |  |  |
| └ Never (base) | 0 (ref.) | — | — | — | 0 (ref.) | — | — | — |
| └ Former | 0.00767 | 0.00358 | 0.00065, 0.01469 | 0.032 | 0.00842 | 0.00365 | 0.00127, 0.01557 | 0.021 |
| └ Current | −0.00807 | 0.00329 | −0.01452, −0.00162 | 0.014 | −0.00782 | 0.00338 | −0.01444, −0.00119 | 0.021 |
| **Self-rated health** |  |  |  |  |  |  |  |  |
| └ Excellent (base) | 0 (ref.) | — | — | — | 0 (ref.) | — | — | — |
| └ Very good | 0.02521 | 0.00647 | 0.01254, 0.03789 | <0.001 | 0.02354 | 0.00672 | 0.01037, 0.03671 | <0.001 |
| └ Good | 0.04126 | 0.00483 | 0.03179, 0.05074 | <0.001 | 0.04016 | 0.00484 | 0.03068, 0.04964 | <0.001 |
| └ Fair | 0.10004 | 0.00511 | 0.09003, 0.11005 | <0.001 | 0.09825 | 0.00512 | 0.08822, 0.10828 | <0.001 |
| └ Poor | 0.16982 | 0.00634 | 0.15739, 0.18224 | <0.001 | 0.16673 | 0.00638 | 0.15423, 0.17923 | <0.001 |
| **Drinking status** |  |  |  |  |  |  |  |  |
| └ Never (base) | 0 (ref.) | — | — | — | 0 (ref.) | — | — | — |
| └ Current drinker | −0.00540 | 0.00292 | −0.01112, 0.00032 | 0.064 | −0.00559 | 0.00295 | −0.01138, 0.00019 | 0.058 |
| **Drinker × Time** | −0.00072 | 0.00033 | −0.00136, −0.00007 | 0.03 | −0.00079 | 0.00033 | −0.00144, −0.00013 | 0.018 |
| **Health insurance** |  |  |  |  |  |  |  |  |
| └ No (base) | 0 (ref.) | — | — | — | 0 (ref.) | — | — | — |
| └ Yes | 0.00095 | 0.00282 | −0.00458, 0.00649 | 0.736 | 0.00121 | 0.00289 | −0.00445, 0.00686 | 0.676 |
| **Cognitive status** |  |  |  |  |  |  |  |  |
| └ Normal (base) | 0 (ref.) | — | — | — | 0 (ref.) | — | — | — |
| └ Instrumental impairment | 0.13899 | 0.01086 | 0.11771, 0.16027 | <0.001 | 0.14615 | 0.01119 | 0.12422, 0.16809 | <0.001 |
| └ CIND | 0.00198 | 0.00295 | −0.00380, 0.00776 | 0.502 | 0.00077 | 0.00292 | −0.00495, 0.00650 | 0.791 |
| └ Dementia | 0.14903 | 0.01381 | 0.12196, 0.17611 | <0.001 | 0.15764 | 0.01373 | 0.13072, 0.18455 | <0.001 |
| **Cognition × Time** |  |  |  |  |  |  |  |  |
| └ Impairment × Time | −0.00571 | 0.00111 | −0.00789, −0.00353 | <0.001 | −0.00580 | 0.00113 | −0.00802, −0.00359 | <0.001 |
| └ CIND × Time | 0.00022 | 0.00034 | −0.00045, 0.00088 | 0.52 | 0.0002 | 0.00034 | −0.00047, 0.00087 | 0.556 |
| └ Dementia × Time | −0.00327 | 0.00151 | −0.00623, −0.00032 | 0.03 | −0.00344 | 0.0015 | −0.00637, −0.00051 | 0.021 |
| **Constant** | 0.06637 | 0.00606 | 0.05449, 0.07825 | <0.001 | 0.06601 | 0.00611 | 0.05404, 0.07798 | <0.001 |
| **Random Effects** |  |  |  |  |  |  |  |  |
| **Women** |  |  |  |  |  |  |  |  |
| └sd(intercept) | 0.0637 | 0.00312 | 0.0579, 0.0700 |  | 0.0624 | 0.0032 | 0.0564, 0.0690 |  |
| └sd(time slope, per year) | 0.00509 | 0.00035 | 0.00444, 0.00583 |  | 0.00515 | 0.00036 | 0.00450, 0.00590 |  |
| └corr(intercept, slope) | 0.249 | 0.15 | −0.0456, 0.543 |  | 0.25 | 0.14 | −0.04, 0.51 |  |
| **Men** |  |  |  |  |  |  |  |  |
| └sd(intercept) | 0.0503 | 0.00247 | 0.0457, 0.0554 |  | 0.0477 | 0.0024 | 0.0431, 0.0527 |  |
| └sd(time slope, per year) | 0.00423 | 0.00036 | 0.00357, 0.00500 |  | 0.00412 | 0.00038 | 0.00344, 0.00494 |  |
| └corr(intercept, slope) | 0.594 | 0.174 | 0.2525, 0.935 |  | 0.62 | 0.14 | 0.28, 0.82 |  |
| **Residual** |  |  |  |  |  |  |  |  |
| └Residual sd (common) | 0.0717 | 0.00109 | 0.0695, 0.0738 |  | 0.0712 | 0.0011 | 0.0690, 0.0734 |  |
| Models were fitted using survey calibration weights with robust standard errors. Multiple imputation estimates were combined using Rubin’s rules. Coefficients (= Coef.) are unstandardised estimates. Age and wealth were centred at the sample mean. Time is measured in years since baseline. Reference (base) group = male, mean age/wealth, never smoker, never drinker, < upper secondary education, non-rural, no vigorous activity, no health insurance, excellent self-rated health, normal cognition. Robust SE = robust standard error. CI = confidence interval. FI = Frailty Index. SRH = self-rated health. CIND = cognitive impairment, no dementia. Impairment = normal with instrumental impairment. | | | | | | | | |

### Supplementary Table S7: Mixed-effects estimates of frailty trajectories on the time-to-death scale among decedents comparing complete-case and multiple imputation analyses.

|  | **Time-to-death Model: Complete case analysis** | | | | **Time-to-death Model: Multiple imputation** | | | |
| --- | --- | --- | --- | --- | --- | --- | --- | --- |
| **Predictor (units; base)** | **Coef.** | **SE** | **95% CI** | **p-value** | **Coef.** | **SE** | **95% CI** | **p-value** |
| **Fixed effects** |  |  |  |  |  |  |  |  |
| **Sex** |  |  |  |  |  |  |  |  |
| **└ Male (base)** | 0 (ref.) | — | — | — | 0 (ref.) | — | — | — |
| **└ Female** | 0.03466 | 0.01032 | 0.01444, 0.05487 | 0.001 | 0.02835 | 0.00982 | 0.00911, 0.04760 | 0.004 |
| **Time (years since baseline)** | −0.03221 | 0.00522 | −0.04245, −0.02198 | <0.001 | -0.03031 | 0.00571 | -0.04149, -0.01912 | <0.001 |
| **Time²** | 0.00237 | 0.00067 | 0.00107, 0.00368 | <0.001 | 0.00219 | 0.00072 | 0.00078, 0.00359 | 0.002 |
| **Time³** | −0.0000586 | 0.000025 | −0.0001076, −0.0000095 | 0.019 | -0.0000538 | 0.0000264 | -0.0001054, -0.0000021 | 0.041 |
| **Female × Time** | −0.00224 | 0.0009 | −0.00400, −0.00048 | 0.013 | -0.00155 | 0.00086 | -0.00324, 0.00014 | 0.072 |
| **Age (centred)** | 0.00106 | 0.00047 | 0.00014, 0.00198 | 0.023 | 0.00139 | 0.00045 | 0.00051, 0.00227 | 0.002 |
| **Age × Time** | −0.00008 | 0.00005 | −0.00017, 0.00001 | 0.083 | -0.000097 | 0.0000435 | -0.000182, -0.0000117 | 0.026 |
| **Wealth (GBP; centred)** | −0.0000000471 | 0.0000000582 | −0.0000001610, 0.0000000671 | 0.419 | -5.35E-08 | 5.79E-08 | -0.000000167, 0.000000060 | 0.355 |
| **Education** |  |  |  |  |  |  |  |  |
| └ < Upper secondary (base) | 0 (ref.) | — | — | — | 0 (ref.) | — | — | — |
| └ Upper secondary/vocational | −0.00269 | 0.01793 | −0.03782, 0.03245 | 0.881 | -0.00372 | 0.0175 | -0.03803, 0.03058 | 0.832 |
| └ Tertiary | −0.02537 | 0.00782 | −0.04070, −0.01005 | 0.001 | -0.02616 | 0.00805 | -0.04194, -0.01039 | 0.001 |
| **Rural residence** |  |  |  |  |  |  |  |  |
| └ No (base) | 0 (ref.) | — | — | — | 0 (ref.) | — | — | — |
| └ Yes | 0.00783 | 0.00533 | −0.00262, 0.01828 | 0.142 | 0.00712 | 0.00518 | -0.00303, 0.01728 | 0.169 |
| **Vigorous activity** |  |  |  |  |  |  |  |  |
| └ No (base) | 0 (ref.) | — | — | — | 0 (ref.) | — | — | — |
| └ Yes | −0.00790 | 0.00539 | −0.01846, 0.00265 | 0.142 | -0.00811 | 0.00526 | -0.01842, 0.00219 | 0.123 |
| **Smoking status** |  |  |  |  |  |  |  |  |
| └ Never (base) | 0 (ref.) | — | — | — | 0 (ref.) | — | — | — |
| └ Former | 0.00189 | 0.00625 | −0.01035, 0.01414 | 0.762 | 0.00227 | 0.00606 | -0.00962, 0.01415 | 0.709 |
| └ Current | −0.02294 | 0.00577 | −0.03425, −0.01163 | <0.001 | -0.02255 | 0.00564 | -0.03359, -0.01150 | <0.001 |
| **Self-rated health** |  |  |  |  |  |  |  |  |
| └ Excellent (base) | 0 (ref.) | — | — | — | 0 (ref.) | — | — | — |
| └ Very good | −0.00174 | 0.01427 | −0.02971, 0.02623 | 0.903 | 0.00072 | 0.01444 | -0.02758, 0.02903 | 0.96 |
| └ Good | 0.02868 | 0.01226 | 0.00465, 0.05271 | 0.019 | 0.02813 | 0.01205 | 0.00451, 0.05176 | 0.02 |
| └ Fair | 0.09019 | 0.01242 | 0.06584, 0.11454 | <0.001 | 0.09051 | 0.01219 | 0.06663, 0.11440 | <0.001 |
| └ Poor | 0.14668 | 0.01316 | 0.12089, 0.17248 | <0.001 | 0.14819 | 0.01287 | 0.12295, 0.17342 | <0.001 |
| **Drinking status** |  |  |  |  |  |  |  |  |
| └ Never (base) | 0 (ref.) | — | — | — | 0 (ref.) | — | — | — |
| └ Current drinker | −0.02427 | 0.0108 | −0.04544, −0.00309 | 0.025 | -0.02598 | 0.01024 | -0.04605, -0.00590 | 0.011 |
| **Drinker × Time** | 0.00175 | 0.00087 | 0.00004, 0.00346 | 0.045 | 0.00189 | 0.00083 | 0.00027, 0.00352 | 0.022 |
| **Health insurance** |  |  |  |  |  |  |  |  |
| └ No (base) | 0 (ref.) | — | — | — | 0 (ref.) | — | — | — |
| └ Yes | −0.00076 | 0.005 | −0.01055, 0.00904 | 0.88 | 0.00124 | 0.00489 | -0.00835, 0.01082 | 0.801 |
| **Cognitive status** |  |  |  |  |  |  |  |  |
| └ Normal (base) | 0 (ref.) | — | — | — | 0 (ref.) | — | — | — |
| └ Instrumental impairment | 0.09719 | 0.0242 | 0.04976, 0.14462 | <0.001 | 0.10177 | 0.02314 | 0.05641, 0.14714 | <0.001 |
| └ CIND | −0.00809 | 0.00994 | −0.02757, 0.01139 | 0.416 | -0.00221 | 0.00998 | -0.02178, 0.01737 | 0.825 |
| └ Dementia | 0.10469 | 0.01926 | 0.06694, 0.14243 | <0.001 | 0.10716 | 0.02039 | 0.06718, 0.14714 | <0.001 |
| **Cognition × Time** |  |  |  |  |  |  |  |  |
| └ Impairment × Time | 0.00322 | 0.00173 | −0.00017, 0.00662 | 0.063 | 0.00205 | 0.00175 | -0.00139, 0.00549 | 0.243 |
| └ CIND × Time | 0.00079 | 0.0009 | −0.00098, 0.00256 | 0.381 | 0.0004 | 0.00089 | -0.00135, 0.00215 | 0.653 |
| └ Dementia × Time | 0.00195 | 0.00276 | −0.00346, 0.00737 | 0.48 | 0.00134 | 0.00276 | -0.00406, 0.00674 | 0.627 |
| **Constant** | 0.2336 | 0.01889 | 0.19657, 0.27063 | <0.001 | 0.22694 | 0.01927 | 0.18918, 0.26469 | <0.001 |
| **Random Effects** |  |  |  |  |  |  |  |  |
| **Women** |  |  |  |  |  |  |  |  |
| └sd(intercept) | 0.1266 | 0.0079 | 0.1121, 0.1431 |  | 0.125 | 0.0077 | 0.110, 0.140 |  |
| └sd(time slope, per year) | 0.00721 | 0.00102 | 0.00546, 0.00951 |  | 0.0667 | 0.0102 | 0.00547, 0.00749 |  |
| └corr(intercept, slope) | −0.93 | 0.03 | −0.97, −0.85 |  | −0.933 | 0.257 | -0.97, -0.82 |  |
| **Men** |  |  |  |  |  |  |  |  |
| └sd(intercept) | 0.1103 | 0.0068 | 0.0976, 0.1245 |  | 0.108 | 0.0069 | 0.096, 0.118 |  |
| └sd(time slope, per year) | 0.00631 | 0.0008 | 0.00492, 0.00809 |  | 0.0582 | 0.0084 | 0.00448, 0.00737 |  |
| └corr(intercept, slope) | −0.98 | 0.03 | −1.00, −0.53 |  | −0.997 | 6.54 | -1.00, 1.00 |  |
| **Residual** |  |  |  |  |  |  |  |  |
| └Residual sd (common) | 0.08235 | 0.00281 | 0.07703, 0.08804 |  | 0.0835 | 0.0028 | 0.081, 0.086 |  |
| Models were fitted using survey calibration weights with robust standard errors. Multiple imputation estimates were combined using Rubin’s rules. Coefficients (= Coef.) are unstandardised estimates. Age and wealth were centred at the sample mean. Time is measured as years before death (time-to-death), with values approaching zero indicating proximity to death. Reference (base) group = male, mean age/wealth, never smoker, never drinker, < upper secondary education, non-rural, no vigorous activity, no health insurance, excellent self-rated health, normal cognition. Robust SE = robust standard error. CI = confidence interval. FI = Frailty Index. SRH = self-rated health. CIND = cognitive impairment, no dementia. Impairment = normal with instrumental impairment. | | | | | | | | |

1. Faculty of Epidemiology and Population Health, London School of Hygiene and Tropical Medicine, London, United Kingdom.
2. School of Health and Care Sciences, University of Lincoln, Leicester, United Kingdom.
